# Epidemiology of Cervical Precancerous Lesions: Prevalence and Predictors from Pap Smear Screening in Hawassa City Hospitals, Sidama Region, Ethiopia. Institutional-Based Cross-sectional Study

**DOI:** 10.64898/2026.06.09.26355254

**Authors:** Azeb Bereket Fisshatsion, Yitateku Alelgn Zewude, Abebe Melis Nisro, Rekiku Fikre Abebe

## Abstract

**Background:** Cervical cancer is the fourth most common cancer in women worldwide and remains a major public health challenge. In Ethiopia, it is the second leading cause of cancer deaths, with around 8,000 new cases and 6,000 deaths each year. Region-specific data on the prevalence and predictors of precancerous lesions remain scarce, yet such information is vital for guiding targeted reproductive health strategies. This study therefore examined the prevalence and predictors of cervical precancerous lesions among women aged 21–60 years undergoing Pap smear screening in public hospitals in Hawassa City, Sidama Region.

**Methods:** An institution-based cross-sectional study was conducted among 241 women attending Pap smear screening at public hospitals in Hawassa City from March to August 2025. Sociodemographic and clinical data were collected via interviews and medical records. Lesions were classified based on the standardized international framework for reporting cervical cytology results from Pap smears per the Bethesda system. Multivariable logistic regression identified predictors p<0.05).

**Result:** Of 241 women screened (mean age 35.3 years), cervical epithelial abnormalities were detected in 52 (prevalence 21.6%). Atypical squamous cells of undetermined significance was the most common abnormality (16.6%). Multivariable analysis showed HIV infection was significantly associated with precancerous lesions (AOR = 3.7, 95% CI: 1.69–8.12, p<0.05), while hormonal contraceptive use was protective (AOR = 0.27, 95% CI: 0.11–0.67, p<0.05).

**Conclusion:** These results underscore the urgent need to strengthen cervical cancer prevention through targeted screening and early intervention. Integrating routine HIV testing with Pap smear programs would be especially valuable. Health authorities should expand accessible screening for women aged 21–60, with particular attention to those living with HIV, to help reduce the burden of precancerous lesions.

## Introduction

Cervical cancer remains a major public health challenge. Globally it ranked the fourth most common cancer among women(1). The burden is disproportionately high in low and middle-income countries, where limited access to preventive and diagnostic services contributes to increased morbidity and mortality(2). According to the Global Cancer Observatory (GLOBOCAN) 2022, an estimated 660,000 new cases of cervical cancer were reported globally, with approximately 94% of the 350,000 deaths occurring in developing countries(3–5). In Ethiopia, cervical cancer is the second most frequent cancer among women after breast cancer and the leading cause of cancer-related death, with 7,745 new cases and 5,338 deaths reported in 2020(6–8). Persistent infection with oncogenic human papillomavirus (HPV) types is recognized as the primary etiological factor(5).

In high-income countries, cervical cancer incidence and mortality have declined significantly due to widespread cytological screening programs, particularly the Papanicolaou (Pap) smear test. This test enables detection of premalignant changes in cervical epithelial cells, allowing timely intervention before progression to invasive cancer(9). Cervical cancer often remains asymptomatic in its early stages. However, advanced disease may present with pelvic pain, abnormal vaginal bleeding, post-coital bleeding, or unusual discharge(10–12).

Precancerous cervical lesions, also referred to as intraepithelial lesions, represent cellular abnormalities that can progress to invasive cancer if left untreated(13,14). Despite being highly preventable through HPV vaccination and cytological screening, cervical cancer continues to cause significant mortality in resource-limited settings due to inadequate screening coverage and treatment facilities(6). In Ethiopia, facility-based study conducted at Debre Markos Referral Hospital reported a prevalence of approximately 14.1% cervical epithelial cell abnormalities among women screened by Pap smear(15).

Recent global studies highlighted an increasing incidence of cervical cancer, driven by population growth, aging, and persistent risk factors such as smoking, immunosuppression, and prolonged use of hormonal contraceptives (9,16). A study conducted in Addis Ababa revealed that many women are diagnosed at advanced stages due to lack of awareness, limited access to quality screening, and inadequate diagnostic and treatment services(17). It is estimated that approximately 33.7 million Ethiopian women aged 15 years and above are at risk of developing cervical cancer(7).

According to the Bethesda System, cervical abnormalities are classified as atypical squamous cells of undetermined significance (ASC-US), atypical squamous cells cannot exclude HSIL (ASC-H), low-grade squamous intraepithelial lesions (LSIL), and high-grade squamous intraepithelial lesions (HSIL). These categories guide management and highlight the importance of early detection(18). Clinical examination alone is insufficient for early detection of cervical precancerous lesions. Cytological screening through Pap smears remains essential for identifying abnormalities at a stage when intervention can prevent progression to invasive cancer(18,19).

Risk factors such as early sexual activity, multiple sexual partners, prolonged use of hormonal contraceptives, smoking, and immunosuppression contribute to disease progression(19). HIV infection, in particular, is strongly associated with increased risk of HPV persistence and cervical epithelial abnormalities, with Ethiopian women living with HIV facing disproportionate morbidity and mortality(20,21).

Implementing Pap smear screening in hospitals and health centers with gynecology and laboratory services provides an opportunity to reduce disease burden and improve treatment outcomes(22). The Pap smear test is simple, cost-effective, and feasible in resource-limited settings, with international guidelines such as those from WHO and American College of Obstetricians and Gynecologists recommending initiation of screening from age 21 years (22–24). Despite the high burden of cervical cancer in Ethiopia, very limited study has assessed the prevalence and predictors of precancerous lesions using Pap smear in Hawassa City. This study determined the prevalence and predictors of cervical epithelial abnormalities among women aged 21–60 years attending public hospitals in Hawassa, Sidama region, Ethiopia,2025.

## Methods

### Study area and design

This institutional-based cross-sectional study was conducted in the Gynecology Outpatient Department and the Pathology Department of two major public hospitals in Hawassa City, Sidama Region, Ethiopia: Adare General Hospital (AGH) and Hawassa University Comprehensive Specialized Hospital (HUCSH). Hawassa, the administrative city of the Sidama Region, is located approximately 275 kilometers south of Addis Ababa.

HUCSH functions as a tertiary referral center, providing specialized medical services for patients from Sidama, Central Ethiopia and Southern Ethiopia regions as well as neighboring zones of the Oromia region. The hospital hosts a well-established Pathology Department that delivers wide range of diagnostic services.

## Study Period

The study was conducted over a six-month period, from March to August 2025.

### Population, Eligibility, Sample size determination and sampling technique and procedure

The study population comprised all women attending gynecological outpatient services HUCSH and AGH in Hawassa City. Inclusion criteria were women aged 21–60 years who presented for gynecological care during the study period and consented to undergo Pap smear screening. Exclusion criteria included women with a prior diagnosis of cervical cancer, those who had undergone hysterectomy, and critically ill patients unable to provide informed consent.

The sample size was determined using the single population proportion formula, assuming a 95% confidence level, 5% margin of error, and a prevalence of cervical epithelial cell abnormalities of 14.1%, based on a facility-based Pap smear study conducted at Debre Markos Referral Hospital(15). This yielded a minimum sample size of 186 participants. After accounting for a 10% non-response rate, the final sample size was 205 women. HUCSH and AGH were selected based on their high patient flow and their established gynecology and pathology services. Eligible women who presented to the gynecology outpatient departments during the study period were subsequently recruited using a systematic sampling technique.

### Study variables, operational definitions

Dependent variable was magnitude of Pap smear detected cervical epithelial cell abnormalities. Whereas the independent variables were socio-demographic factors (age, marital status, occupation, educational status), sexual and reproductive history (number of sexual partners, age at first sexual intercourse, number of children), clinical factors (HIV status, STI history), and behavioral factors (smoking, hormonal contraceptive use).

#### Precancerous cervical lesion

Defined as the presence of epithelial cell abnormalities detected on Pap smear cytology, specifically Atypical Squamous Cells of Undetermined Significance (ASC-US), Atypical Squamous Cells cannot exclude HSIL (ASC-H), Low-grade Squamous Intraepithelial Lesion (LSIL), and High-grade Squamous Intraepithelial Lesion (HSIL), classified according to the Bethesda System 2014.

### Data collection and procedures

Data were collected by trained gynecology residents and nurses using two approaches: Pap smear specimen collection and structured interviews. Pap smear samples were obtained after explaining the study objectives and securing oral informed consent. Smears were fixed with 95% ethyl alcohol and processed in the pathology laboratory using the Papanicolaou staining method. Microscopic examination was performed by pathology residents and pathologists, and results were interpreted according to the Bethesda System 2014. Socio-demographic and clinical data were collected via face-to-face interviews using a structured, pretested questionnaire developed from relevant literature.

To ensure data quality, a one-day training was provided to data collectors and sample collectors covering study objectives, Pap smear collection techniques, questionnaire administration, and confidentiality. Close supervision was maintained throughout the data collection period, with daily checks for completeness and consistency. Proper coding, categorization, and verification against patient records were performed before data entry.

### Data Analysis

Data were collected using Kobo Toolbox and exported to Stata version 17 for analysis. Cleaning was performed to check for missing values and inconsistencies. Descriptive statistics (frequencies, percentages, means, and standard deviations) were used to summarize variables. Associations between cervical epithelial abnormalities and categorical variables were assessed using chi-square tests. Bivariable and multivariable binary logistic regression analyses were conducted to identify independent predictors, with variables having p-values < 0.25 in bivariable analysis included in the multivariable model. Model fitness was assessed using the Hosmer-Lemeshow test, and odds ratios (ORs) with 95% confidence intervals (CIs) were reported. A *p* value < 0.05 was used to estimate the strength and significance of the association.

### Ethical Considerations

Ethical clearance was obtained from the Institutional Review Board (IRB) of Hawassa University College of Medicine and Health Sciences (Ref. No. IRB/440/17). Formal letters of support were submitted to each participating hospital. Written informed consent was obtained from all participants, with the consent form translated into the local language and read aloud when necessary. Confidentiality was strictly maintained, and participants were informed of the study objectives, their right to withdraw at any time, and the respect for their privacy and dignity throughout the study. No identifiable personal information was retained in the dataset used for analysis.

## Result

### Sociodemographic characteristics of participants

This study included 253 women, of whom 12 were excluded because their samples were inadequate and the final analysis was conducted on 241 cases. The mean age of the participants was 35.3 years (SD = 8.3; range 21–60). In terms of age categories, 11.2% of participants were older than 45 years. The majority (75.9%) were married, and 85.5% resided in urban areas. Nearly one-quarter (24.1%) had completed college or above, while an equal proportion (24.1%) had no formal education. More than half of the participants (53.9%) were housewives. (Table 1)

**Table 1.**
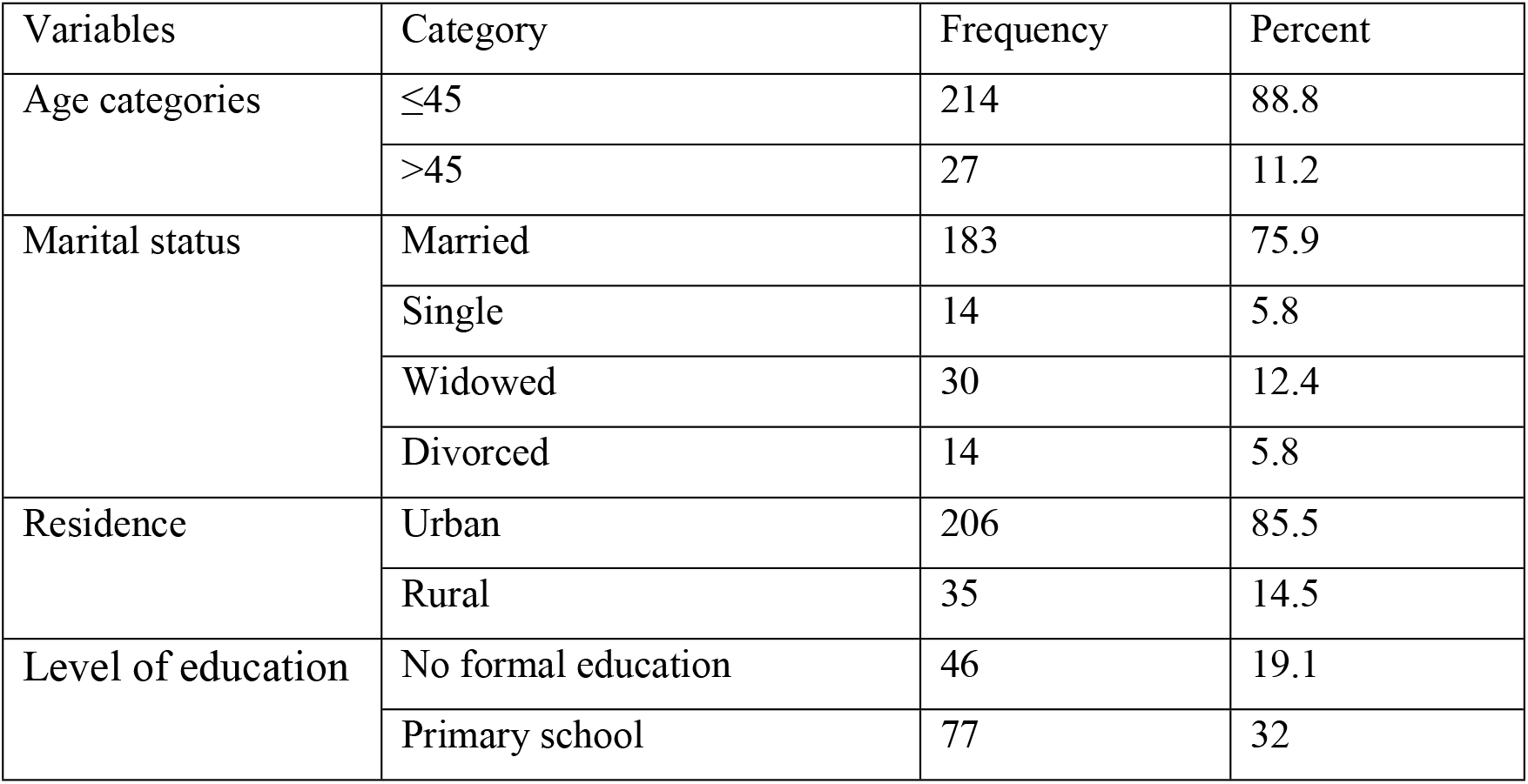
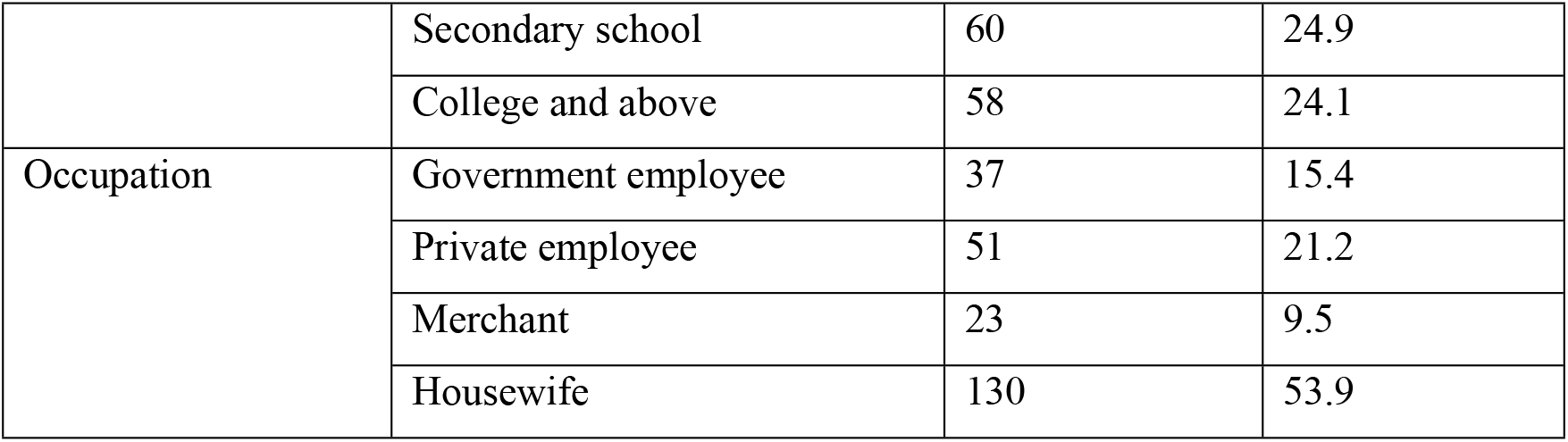
Sociodemographic characteristics of study participants.

### Behavioral, reproductive and clinical characteristics of the participants

Regarding sexual behavior, 30% of women reported having multiple sexual partners, and 31.1% reported a history of sexually transmitted infections. Overall, 25.3% of participants were HIV positive. The median age at first sexual intercourse was 18 years (IQR: 16–20). Family history of cervical cancer was reported by 5.8% of the participants. A total of 216 women (89.6%) reported a history of pregnancy. The median age at first pregnancy was 20 years, and the median parity was three (IQR: 2–4). Hormonal contraceptive use was reported by 37.3%. The mean age at menarche was 14.3 years (SD = 1.2). Most participants (90%) delivered by spontaneous vaginal delivery (SVD). (Table 2)

**Table 2.** Behavioral, reproductive and clinical characteristics of study participants.

| Variables | Category | Frequency | Percent |
| --- | --- | --- | --- |
| Life time sexual partners | Single | 169 | 70 |
|  | Multiple | 72 | 30 |
| History of STI | Yes | 75 | 31.1 |
|  | No | 166 | 68.9 |
| HIV status | Positive | 61 | 25.3 |
|  | Negative | 180 | 74.7 |
| Family history of cervical cancer | Yes | 14 | 5.8 |
|  | No | 227 | 94.2 |
| History of pregnancy | Yes | 216 | 89.6 |
|  | No | 25 | 10.4 |
| Hormonal contraceptive use | Yes | 90 | 37.3 |
|  | No | 151 | 62.7 |
| Mode of delivery | SVD | 190 | 90 |
|  | C/S | 21 | 10 |

### Cervical Screening Results

Visual inspection with acetic acid (VIA) was performed for 232 women, of whom 34 (14.7%) tested positive. Pap smear examination was done and reported based on the Bethesda 2014 classification for 241 women. Overall, 52 (21.6% CI: 16.8-27.2%) had cervical epithelial abnormalities, while 189 (78.4%) were NILM. Among the abnormalities, ASC-US were most frequent, detected in 16.6%. (Figure 1)

**Figure 1.**
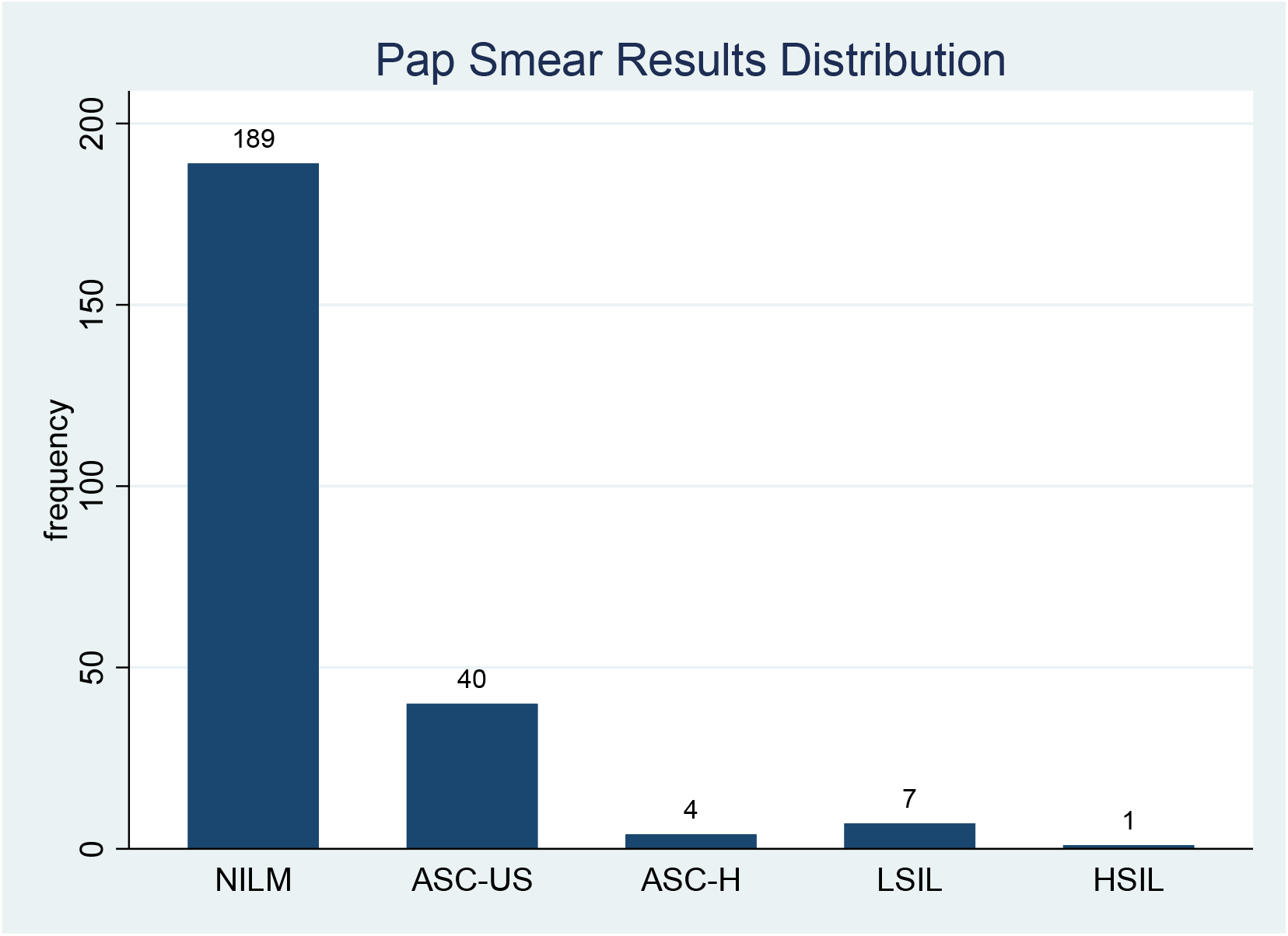
Pap smear result distribution NILM=Negative for Intraepithelial Lesion or Malignancy, ASC-US=Atypical Squamous Cells of Undetermined Significance, ASC-H=Atypical Squamous Cells, cannot exclude High-grade Squamous Intraepithelial Lesion, LSIL=Low-grade Squamous Intraepithelial Lesion, HSIL=High-grade Squamous Intraepithelial Lesion.

### Predictors of cervical precancerous lesions

In bivariable analysis, age, educational level, number of lifetime sexual partners, HIV status, and contraceptive use showed statistically significant associations with precancerous cervical lesions.

However, in the multivariable logistic regression model, only HIV status and contraceptive use remained independently associated. HIV-positive women had 3.7 times higher odds of precancerous cervical lesions compared to HIV-negative women (AOR = 3.7, 95% CI: 1.69–8.12, p = 0.001). In contrast, contraceptive use was protective, with users having 73% lower odds of precancerous cervical lesions compared to non-users (AOR = 0.27, 95% CI: 0.11–0.67, p = 0.005) (Table 3).

**Table 3:**
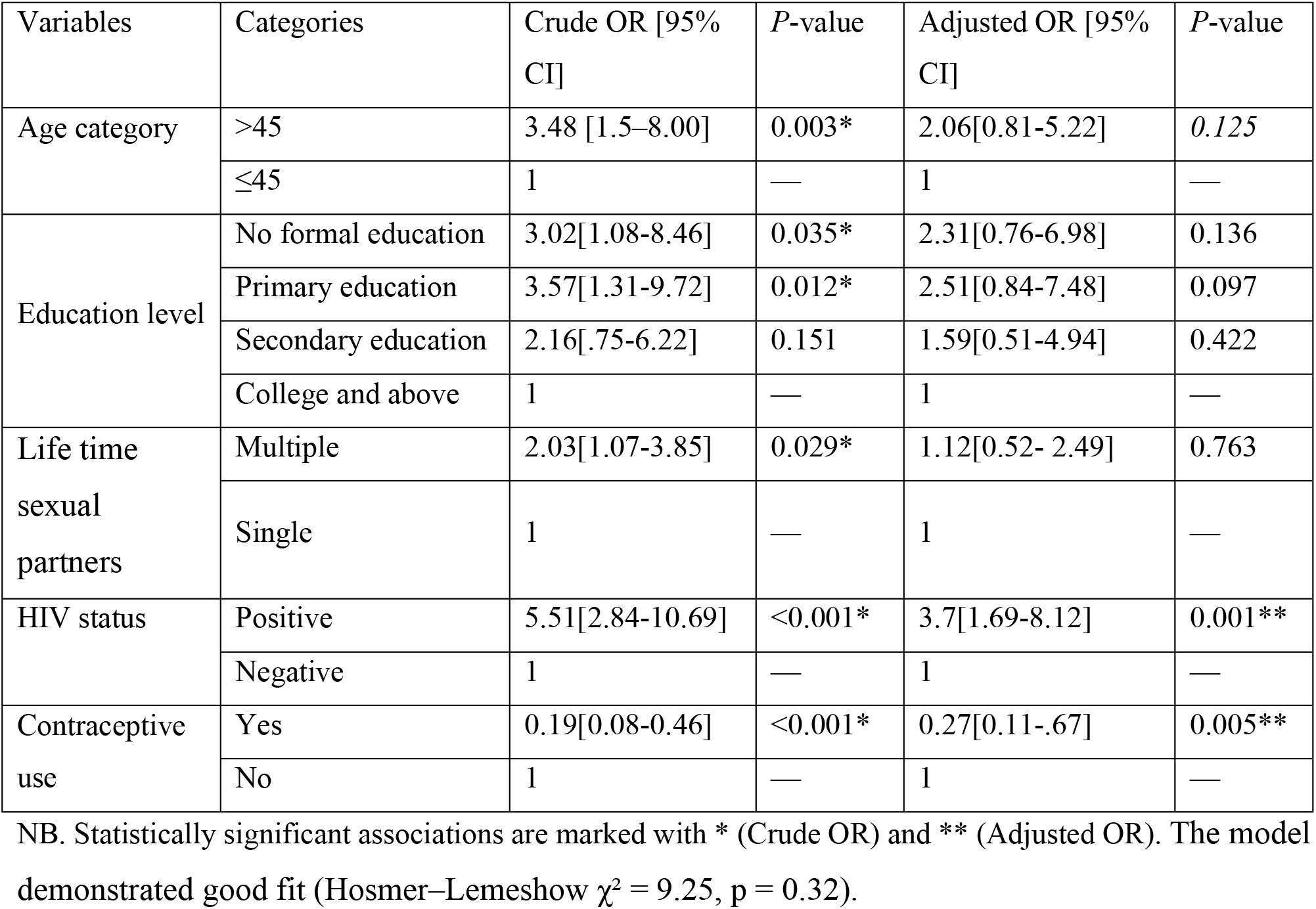
Bivariable and Multivariable Predictors of cervical precancerous lesions (n=241)

## Discussion

This study showed magnitude and predictors of cervical precancerous lesions among women screened by Pap smear in public hospitals of Hawassa City. The prevalence of epithelial abnormalities was 21.6%, which is comparable to the 18% prevalence reported in southern Ethiopia study(23). However, this finding is higher than most national and international reports. For example, a study conducted at Debre Markos Referral Hospital documented a prevalence of 14.1%, while a systematic review and meta-analysis on precancerous cervical lesions among Ethiopian women reported a pooled prevalence of 14.4% (15,25). This relatively higher prevalence in our study may be explained by the hospital-based nature of the sample, where women are more likely to present with symptoms or be referred for evaluation, unlike community-based screening programs.

In the multivariable analysis, HIV infection and contraceptive use were independently associated with cervical precancerous lesions. Women living with HIV had 3.7 times higher odds of developing epithelial abnormalities compared to HIV-negative women. This finding is consistent with previous Ethiopian studies(15,20). and international evidence linking immunosuppression to persistent HPV infection and progression to cervical intraepithelial neoplasia(21). The elevated risk among HIV-positive women can be explained by the immune suppression that facilitates persistent HPV infection, reduced clearance of oncogenic strains, and the synergistic effect of co-existing sexually transmitted infections. The high burden of cervical cancer among women living with HIV has been well documented, and global initiatives emphasize the integration of cervical cancer screening into HIV care services(6).

Conversely, hormonal contraceptive use appeared protective in our study, with users showing 73% lower odds of precancerous lesions. Although some studies have suggested that prolonged hormonal contraceptive use may increase the risk of cervical cancer through hormonal influences on cervical epithelium and HPV persistence(9,16), other evidence emphasizes the role of healthcare engagement. Women who use contraceptives are often more likely to access reproductive health services, which increases opportunities for screening, counseling, and early detection of cervical abnormalities(17,24). Our finding is consistent with reports from Bahir Dar(23) and Addis Ababa(17), supporting the interpretation that in resource-limited settings, contraceptive use may function less as a direct biological protective factor and more as a proxy for improved healthcare access and utilization.

## Conclusion

This study found that cervical precancerous lesions is a public health concern in Hawassa City, with HIV infection significantly increasing risk and contraceptive use showing a protective association. The elevated prevalence likely reflects the hospital-based nature of the cohort, underscoring the need for targeted interventions in high-risk populations. These findings emphasize the importance of integrating cervical cancer screening into HIV and reproductive health services, while also strengthening national strategies such as HPV vaccination and awareness programs. Further studies are needed to clarify the protective role of contraceptive use, especially in resource-limited settings.

## Strengths and limitations of the study

A major strength of this study is the use of Pap smear cytology interpreted according to the Bethesda System, ensuring standardized reporting. The inclusion of both sociodemographic and clinical predictors provides a comprehensive analysis. However, limitations include the cross-sectional design, which precludes causal inference, and reliance on self-reported behavioral data, which may be subject to recall bias. Additionally, the study was facility-based, which may limit generalizability to the wider population.

## Data Availability

The datasets used or analyzed during the current study are available from the Hawassa University College of Medicine and Helath Sciences IRB and corresponding author.

## Acknowledgment

We would like to express our sincere gratitude to the gynecology residents and nurses, as well as to all staff members of the pathology departments, particularly the pathologists and residents, for their invaluable support.

## Authors contribution

A.B., Y.A., A.M. and R.F. design methodology, collect the data, analysis the data, and write up the result and manuscript.

